# Risk Perceptions, Knowledge and Behaviors of General and High-Risk Adult Populations towards COVID-19: A Systematic Scoping Review

**DOI:** 10.1101/2021.02.09.21250257

**Authors:** Nathalie Clavel, Janine Badr, Lara Gautier, Mélanie Lavoie-Tremblay

**Author notes:** **Corresponding author:** Nathalie Clavel, PhD, Postdoctoral Fellow, Ingram School of Nursing, McGill University, 680, Sherbrooke West Street, 18^th^ floor, Montreal, H3A 2M7, Canada.

## Abstract

**Background:** The COVID-19 pandemic represents a major crisis for governments and populations around the globe. A large number of studies have been conducted worldwide to understand people’s awareness and behavioral response towards the disease. The public’s risk perceptions, knowledge, and behaviors are key factors that play a vital role in the transmission of infectious diseases. Our scoping review aims to map the early evidence on risk perceptions, knowledge, and behaviors of general and high-risk adult populations towards COVID-19.

**Methods:** A systematic scoping review was conducted of peer-reviewed articles in five databases (MEDLINE-Ovid, EMBASE-Ovid, PsycINFO-Ovid, Web of Science, and CINAHL-EBSCO) on studies conducted during the early stage of COVID-19 (January to June, 2020). The gray literature was also searched through Open Grey, Scopus, Wonder, Social Science Research Network, MedRxiv, and websites of major public health organizations. Twenty studies meeting the inclusion criteria were included, appraised and analyzed.

**Results:** During the early stage of the pandemic, levels of risk perceptions, knowledge, and behaviors towards COVID-19 were moderate to high in both general and high-risk adult populations. The perceived severity of the disease was slightly higher than the perceived susceptibility of getting COVID-19 during the first wave of COVID-19. Adults were knowledgeable about preventive behaviors, including hand-washing, mask-wearing, social distancing, and avoidance behaviors. Nevertheless, an important knowledge gap regarding the asymptomatic transmission of COVID-19 was reported in many studies. Our review identified hand-washing and avoiding crowded places as dominant preventive behaviors at the early stage of the pandemic. Staying at home, reducing social contacts, and avoiding public transport were less widespread in general populations than in high-risk adult groups. Being a female, older, and more educated was associated with better knowledge of COVID-19 and appropriate preventive behaviors.

**Conclusion:** This scoping review offers a first understanding of general and high-risk adults’ risk perceptions, knowledge, and behaviors towards COVID-19 during the early stage of the COVID-19 pandemic. Further research should be undertaken to assess psychological and behavioral responses over time. Research gaps have been identified in the relationship between ethnicity and risk perceptions, knowledge, and behaviors towards COVID-19.

**Contribution to the field statement:** Since the beginning of the pandemic, a large number of primary studies have been conducted worldwide to understand people’s awareness and behavioral response towards COVID-19. Nevertheless, no review has mapped the early evidence on the perceptions, knowledge, and preventive behaviors of adult populations towards the transmission of this new disease. To the best of our knowledge, this is the first scoping review that offers an understanding of the general and high-risk adults’ risk perceptions, knowledge, and behaviors (RPKB) towards COVID-19 during the early stage of the COVID-19 pandemic. This review also identified sociodemographic factors associated with adults’ RPKB regarding COVID-19. As the virus does not affect individuals equally, knowing these factors can help to mitigate the negative effects of COVID-19 in certain population groups by developing targeted communication strategies that will facilitate their engagement in preventive measures. Finally, research gaps have been identified in the relationship between ethnicity and RPKB towards COVID-19. The existence of a disproportionate number of COVID-19 fatalities within Black populations should signal the possible gaps in RPKB towards COVID-19 in these communities. Additional studies on ethnic health disparities can help public health authorities to introduce targeted actions towards these communities during the COVID-19 pandemic.

## Background

Coronavirus disease (COVID-19) was declared a pandemic by the World Health Organization on March 11, 2020. Since then, COVID-19 continues to represent a major concern for populations and governments. As of mid-January, 2021, more than 92 million cases have been confirmed and 2 million confirmed COVID-19-related deaths have been reported globally (1). Among the adult populations, older people and individuals with underlying health conditions are at the greatest risk for developing severe complications (2). An increasing number of authors suggest that individuals of disadvantaged socioeconomic groups, because of their greater exposure to the virus, have an increased risk of infection with COVID-19 (3, 4). Similarly, evidence suggests that some ethnic minority groups are at increased risk of getting sick and dying from COVID-19 (4-6).

Since the beginning of the COVID-19 pandemic, a large number of studies have been conducted worldwide to understand people’s awareness and behavioral response towards the disease. Public risk perceptions (RPs), knowledge, and behaviors are key factors that play a vital role in the community transmission of infectious diseases (7, 8). Previous coronavirus outbreaks, like Severe Acute Respiratory Syndrome (SARS-CoV-1) in 2003 and the Middle East respiratory syndrome (MERS) in 2012, have shown that public awareness and compliance with preventive measures play key roles in controlling the spread of the virus (9, 10). Several behavior change models have been applied to assess public response to infectious outbreaks (11). One of the most widely used in public health is the knowledge, attitude, and practice (KAP) model that identifies gaps in KAP in relation to health (12). KAP studies conducted during past infectious disease outbreaks generally assessed various aspects of knowledge (e.g., routes of transmission, common symptoms, preventive behaviors), attitudes (e.g., risk perceptions, impact on daily life) and preventive practices (avoidance behaviors, mask wearing, social distancing) (9, 10, 13). The health belief model, another widely used health-related behavioral model, argues that RPs, including perceived susceptibility and perceived severity of a disease, are key contributors to people’s behavior changes during pandemics (11). Behavioral change models rely on the hypothesis that knowledge and/or attitudes (public perceptions, including risk perceptions) are predictors of the behaviors of individuals (11, 14). Thus, simultaneously identifying RPs, knowledge, and behaviors (RPKB) of general and high-risk adult populations can inform risk communication strategies and interventions to better control the spread of COVID-19. As the virus does not affect individuals equally, the factors associated with RPKB that can explain why certain population groups are more likely to get infected with COVID-19 must be identified. Knowing these factors can help mitigate the negative effects of COVID-19 in high-risk groups as more targeted strategies can be developed to facilitate engagement in the preventive measures.

To the best of our knowledge, no overviews have been published of primary studies assessing RPKB of general and high-risk adult populations with regards to COVID-19. The objectives of our scoping review were therefore to 1) conduct a systematic search of the recently published primary studies assessing RPKB of general and high-risk adult populations with regards to COVID-19; 2) map the characteristics of the identified studies; and 3) identify the level of RPKB towards COVID-19 in these populations and the factors associated with RPKB.

## Methods

Given the high number of studies conducted with only short delays, we decided to conduct a scoping review to map the early evidence regarding our research questions (15). To ensure a systematic approach in conducting our scoping review and for reporting the findings, we followed the PRISMA extension for scoping reviews (PRISMA-ScR) (16). Before conducting the review, we published a research protocol on the protocols IO research platform (17). We conducted a comprehensive search of the following electronic databases: MEDLINE-Ovid, EMBASE-Ovid, PsycINFO-Ovid, Web of Science, and CINAHL (EBSCO). The searches were performed in English, with the search terms in Table 1.

**Table 1:** Search terms.

| Population | Perception | Knowledge | Behavior |
| --- | --- | --- | --- |
| Public | Perception<br>Risk perception | Knowledge | Behavior |
| People | Awareness | Comprehension | Practice |
| Person | Consciousness |  | Action |
| Individual |  |  |  |
| Resident |  |  |  |
| Citizen |  |  |  |
| Adult |  |  |  |
| Community |  |  |  |
| Group |  |  |  |
| Patient |  |  |  |

The search term strategies were developed with support from a librarian at X University. Our search strategy for MEDLINE-Ovid is shown in Appendix 1. Searches using the four other databases are available upon request. A comprehensive search of the gray literature was undertaken through Open Grey, Scopus, Wonder, Social Science Research Network, and MedRxiv. We also searched the World Health Organization, Centers for Disease Control and Prevention, European Centre for Disease Prevention and Control, and the Center for Infectious Disease Research and Policy websites.

### Inclusion and exclusion criteria

We included peer-reviewed and preprint articles that assessed RPKB of general adult populations or high-risk adults with regards to COVID-19. High-risk groups were defined based on the Centers for Disease Control and Prevention’s definition (18) as individuals who are at greater risk of getting infected with COVID-19 or developing severe illness from SARS-CoV-2 because of their age, underlying health conditions, or socio-economic conditions. We decided to exclude studies that did not simultaneously assess RPKB related to COVID-19, because all of the factors in combination play a role in the transmission of the virus. The inclusion and exclusion criteria are shown in Table 2.

**Table 2:** Inclusion and exclusion criteria.

| Inclusion criteria | Exclusion criteria |
| --- | --- |
| Peer reviewed or preprint articles | Studies not based on original research (e.g., editorial, opinion, or commentary papers) |
| Risk perceptions, knowledge and behaviors towards COVID-19 | Studies that did not simultaneously assess risk perceptions, knowledge, and behaviors towards COVID-19 |
| Adults | Children or adolescents |
| General or high-risk populations | Healthcare workers or students in medicine, dentistry or health sciences (e.g., nursing) |
| Any study design | Studies using data from online posts or searches (e.g., data from Google searches) |
| English language |  |
| Published or posted between January and June 2020 |  |

### Data screening and extraction

Two authors (NC and JB) independently reviewed the titles and abstracts of the articles against our inclusion and exclusion criteria. A pilot round with a randomly generated sample of nearly 10% of the articles was done to evaluate inter-reviewer agreement on the exclusion and inclusion criteria before a full screening was done for all articles (19, 20). After the initial review of full-text articles, four authors (NC, JB, LG, MLT) completed the data extraction. The data extraction consisted of collecting variables on the 1) general characteristics of the articles (i.e., authors, title, month of publication, country, and publication journal); 2) characteristics of the participants (i.e., data collection period and method, targeted population, sample size and characteristics, sampling scheme, response rate, and statistical analysis); and 3) research question outcomes (i.e., RPKB in general and high-risk adult populations and their associated factors).

### Quality appraisal

In the context of the COVID-19 pandemic where studies have been conducted in very short time-frames, we decided to assess the quality of the included articles. Since all articles were cross-sectional studies, the quality of studies was assessed using the appraisal tool for cross-sectional studies (AXIS tool) (21), which was developed to appraise observational cross-sectional studies (21). Quality appraisal was conducted independently by the four co-authors (NC, JB, LG, and MLT). Discrepancies were resolved through discussions between the first author (NC) and the co-authors (JB, LG, and MLT). A total score was assigned to each article, which was then grouped into one of three categories: 1) low quality (<60%), 2) medium quality (60-80%), and 3) high quality (≥80%). Low quality studies were not excluded from the scoping review.

## Results

Our initial search yielded 4,878 articles and 60 additional records were identified through the preprint servers. After removing duplicates and reviewing the titles and abstracts, 20 articles met our inclusion criteria (see the PRISMA flow diagram in Figure 1 and Table 3).

**Table 3:**
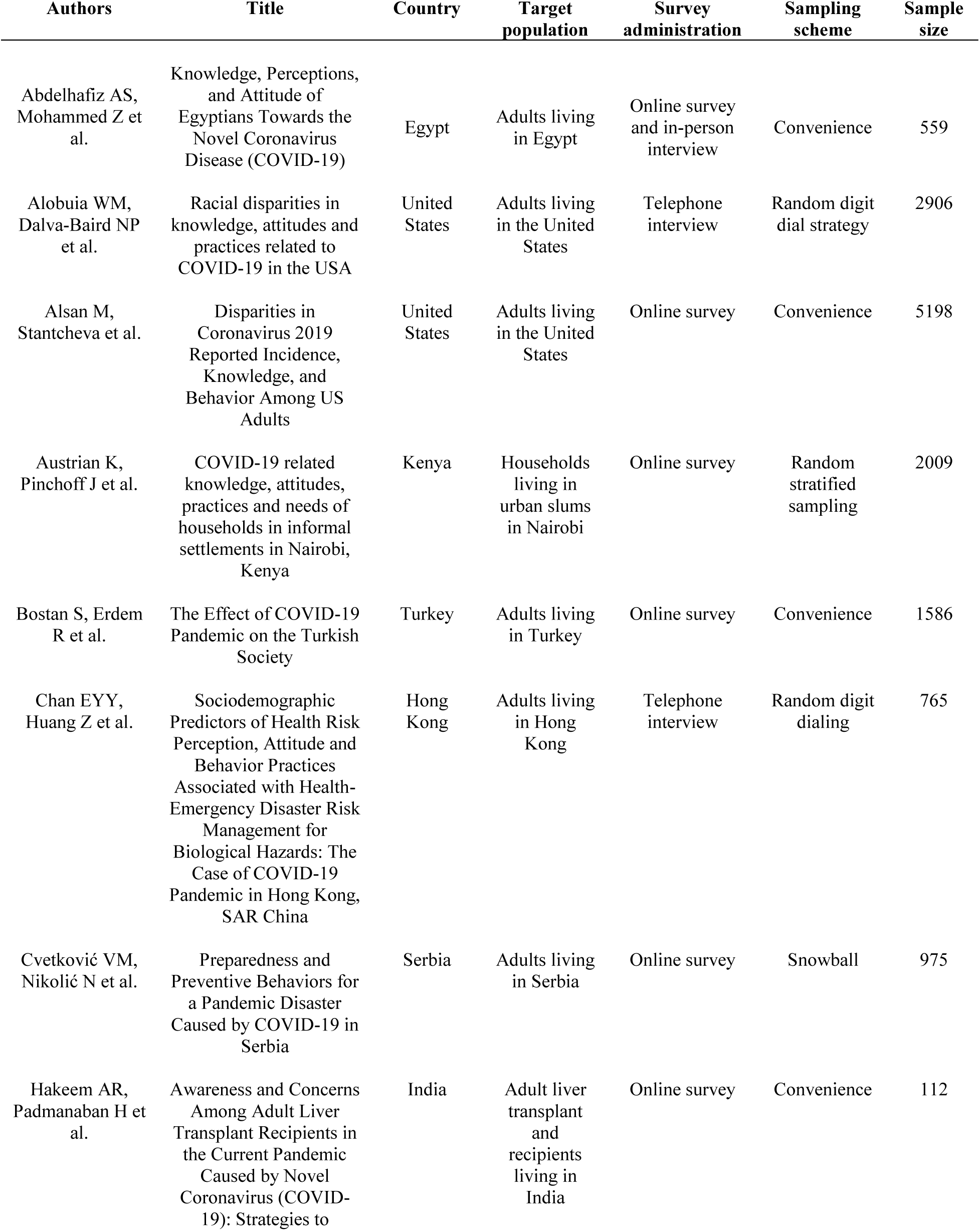
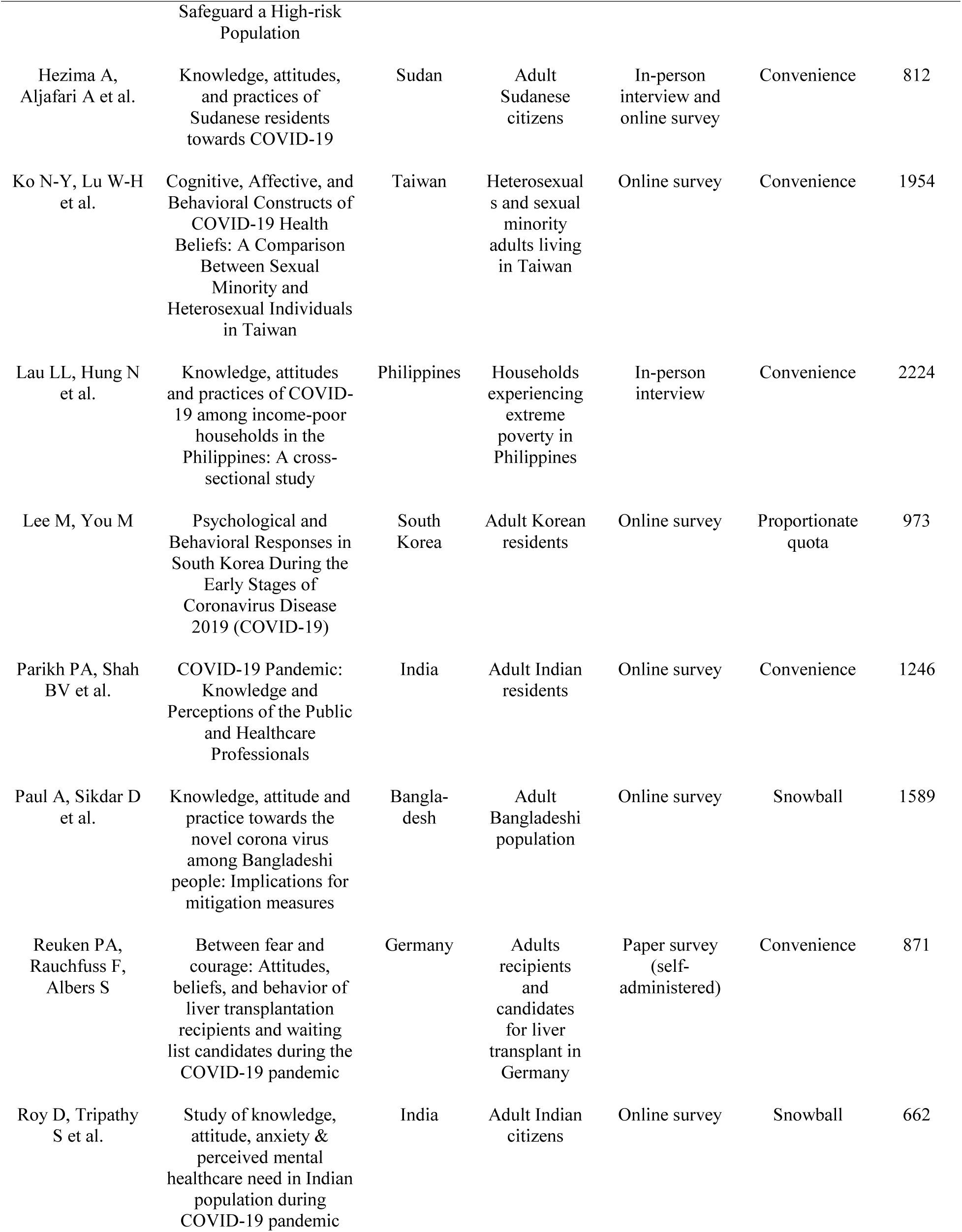
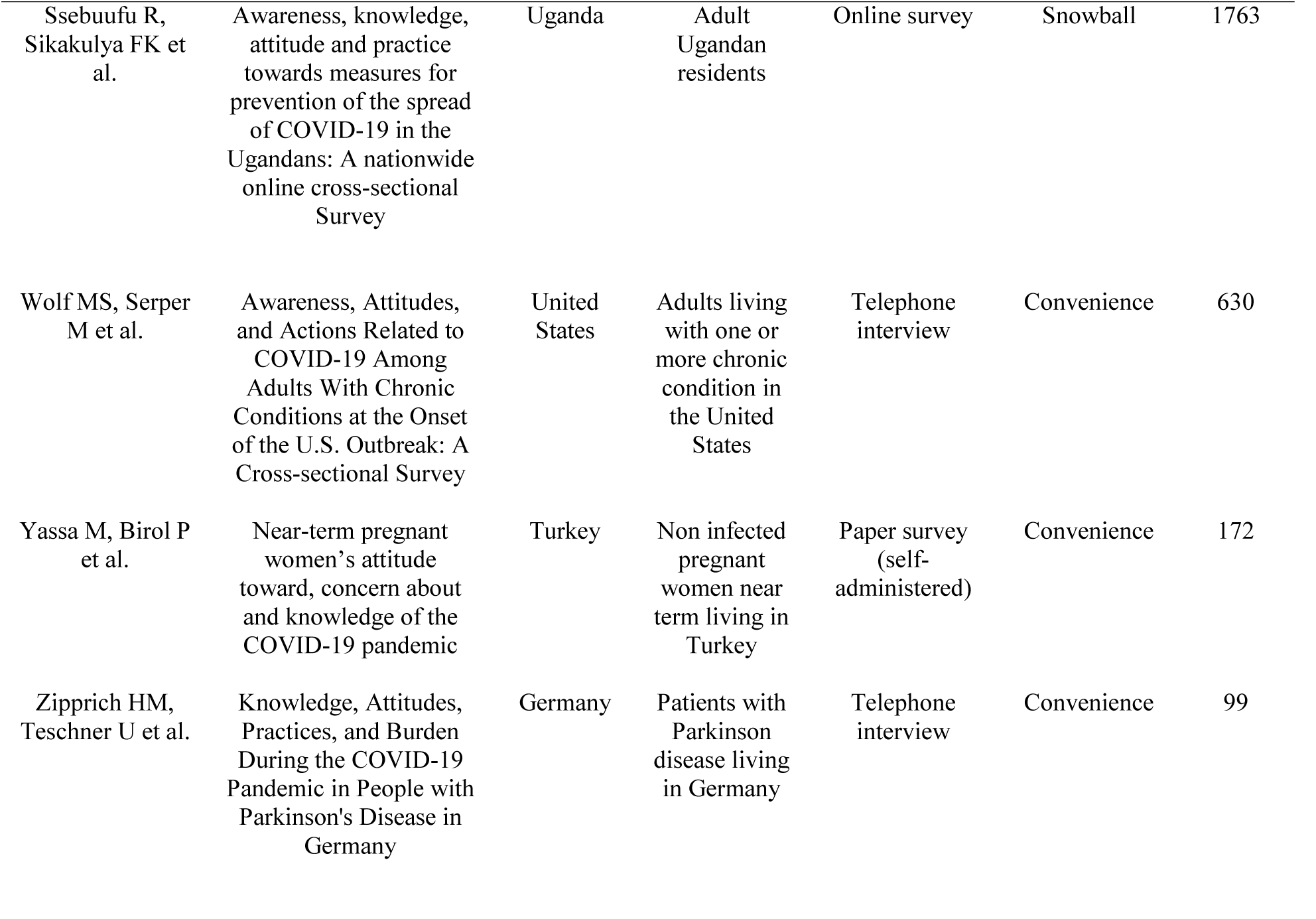
Articles assessing risk perceptions, knowledge and behaviors towards COVID-19.

### General characteristics of the included studies

All of the studies were cross-sectional, using surveys conducted between February and April 2020. Most of the studies (75%) were based on non-probability sampling methods while only 20% used random sampling methods. A total of 55% of the studies had samples that were smaller than 1000 participants. Finally, 80% of the articles used both descriptive and advanced statistics to present the survey data and analyze factors that were significantly associated with RPKB towards COVID-19. The main characteristics of the included studies are shown in Table 4.

**Table 4:** General characteristics of the included studies (n=20)

|  | Number<br>(%) of<br>studies |
| --- | --- |
| **Study design** |  |
| Cross-sectional (surveys) | 20 (100) |
| **Sample size** |  |
| <500 | 3 (15) |
| ≥500-999 | 8 (40) |
| ≥1000-1999 | 5 (25) |
| ≥2000 | 4 (20) |
| **Sampling scheme** |  |
| Convenience sampling | 11 (55) |
| Random and stratified sampling | 4 (20) |
| Snowball sampling | 4 (20) |
| Quota sampling | 1 (5) |
| **Data collection period** |  |
| February | 2 (10) |
| March | 13 (65) |
| April | 5 (25) |
| **Use or adaptation of an existing scale/survey** |  |
| Yes | 5 (25) |
| No | 15 (75) |
| **Data collection method/mode of administration\*** |  |
| Online survey (self-administered) | 12 (60) |
| Phone or face-to-face (administered by an interviewer) | 8 (40) |
| Paper survey (self-administered) | 2 (10) |
| **Statistical analysis** |  |
| Descriptive statistics (only) | 4 (20) |
| Descriptive and advanced statistics<br>(analysis of variance/regression analysis) | 16 (80) |

\* The total sometimes exceeds 100%, because two studies used two survey administration methods for the participants.

### Characteristics of the participants

The survey participants were from 14 countries across four continents (Africa, Asia, Europe, and North America). Most of the studies assessed RPKB towards COVID-19 among general adult populations (n=12), while eight studies focused on high-risk adults. All studies collected demographic statistics on age and gender and most of them collected data on the level of education and income. Other data were related to living areas, occupation, health status, and ethnicity. More detailed information is provided in Table 5.

**Table 5:** Characteristics of participants.

|  | Number (%) of studies |
| --- | --- |
| **Country of residence** |  |
| United-States | 3 (15) |
| India | 3 (15) |
| Germany | 2 (10) |
| Turkey | 2 (10) |
| Bangladesh | 1 (5) |
| Hong Kong | 1 (5) |
| Egypt | 1 (5) |
| Kenya | 1 (5) |
| Korea | 1 (5) |
| Philippines | 1 (5) |
| Serbia | 1 (5) |
| Sudan | 1 (5) |
| Taiwan | 1 (5) |
| Uganda | 1 (5) |
| **Population targeted** |  |
| General adult population | 12 (60) |
| Adults with a chronic condition | 4 (20) |
| Poor households | 2 (10) |
| Pregnant women | 1 (5) |
| Sexual minorities | 1 (5) |
| **Demographic and other statistics collected\*** |  |
| Age | 20 (100) |
| Gender | 20 (100) |
| Level of education | 15 (75) |
| Income | 10 (50) |
| Living areas | 9 (45) |
| Occupation | 8 (40) |
| Health status or chronic condition | 5 (25) |
| Ethnicity | 3 (15) |

\* The total sometimes exceeds 100%, because some studies collected different types of statistics for the participants.

### Quality appraisal of the included studies

Overall, the quality of the included studies was quite low. The mean score calculated using the AXIS quality appraisal tool was 61%. Most studies were of low quality (n=9) or medium quality (n=9) and only two were of high quality. Low-quality articles were mostly of studies conducted in general adult populations that used online surveys and convenience sampling or snowball methods to recruit participants without accounting for possible selection and non-response bias, which led to sampling bias with young or educated participants being overrepresented in the samples. These studies usually did not report the response rate and did not provide any information about non-responders, which can lead to non-response bias and over- or under-representation of certain categories of the populations. Finally, most of the articles (n=15) did not report using or adapting existing tools to assess RPKB related to COVID-19. The detailed quality appraisal grid is shown in Appendix 2. The low quality of the included articles implies that the results should be interpreted carefully, especially for the lowest-rated studies.

### RPKB in general and high-risk adult populations

#### Risk perceptions towards COVID-19

In 85% of the studies (n=17), RPs towards COVID-19 were assessed through perceived susceptibility for self and/or others to be infected. Only one study also measured risk perception by considering the concept of the perceived risk of infecting others (22). Perceived severity of COVID-19 in the community or for high-risk groups was assessed in 65% of the studies (n=13). RPs were reported by the percentage of participants who reported being worried about getting infected or by calculating the mean score of the perceived likelihood of becoming infected (low or high scores).

#### Perceived susceptibility towards COVID-19

##### General adult populations

Overall, the studies found that perceived susceptibility for self was moderate to high in the population. Three studies conducted in the United States (23), Hong Kong (24) and India (25) reported a moderate proportion of adults being worried about getting infected (ranging from 40% to 67%). Two studies in Egypt (26) and India (27) showed a higher proportion of participants (87% and 82 % respectively) being worried about getting infected. Among studies that measured the perceived likelihood of getting sick from COVID-19 among adults in the United States (28), South Korea (29) and Serbia (30), the perceived risk was moderate (mean scores ranged from 3.9/10 to 3/5) (28-30).

##### High-risk adults

Most of the studies conducted among high-risk adults also found a high level of perceived susceptibility towards COVID-19. Four studies reported a high percentage of participants being worried about getting infected (ranging from 87% to 64%). These studies were conducted among adults living with a chronic disease in the United States (31), poor households in the Philippines (32), sexual minorities in Taiwan (33) or liver transplantation recipients and candidates for transplants in Germany (34). In addition, two studies reported a low percentage of perceived susceptibility among the participants (35%); one was conducted among pregnant women in Turkey (35) and the other was in poor households in Kenya (36).

#### Perceived severity of COVID-19

##### General adult populations

The perceived severity of COVID-19 among the participants was high in most studies that measured this variable (n=5). The proportion of participants who perceived COVID-19 as a serious threat to their health ranged from 97% to 80%, in studies in Hong Kong (24), Sudan (37), and Uganda (38). We noted the exception of one study that was conducted in Bangladesh where 55% of the participants considered COVID-19 as a deadly disease (39). In studies that measured perceived severity, the mean scores among participants in South Korea (29) and Serbia (30) ranged from 3.66/5 to 4.7/5 (29, 30).

##### High-risk adults

As in the general adult populations, the perceived severity of the disease among high-risk adults was significant. Four studies reported a high proportion of participants who perceived COVID-19 (68% to 95%) as a serious threat for themselves. The studies were conducted among liver recipients and candidates for transplants (34), adults with Parkinson’s Disease (40), sexual minorities (33), and poor households (36). In the study conducted among pregnant women, only 51% of the women felt more vulnerable to developing complications from COVID-19 (35).

#### Knowledge of COVID-19

Knowledge related to COVID-19 was mainly assessed through five main variables: modes of transmission (n=12), common symptoms (n=12), preventive behaviors to avoid infection (n=9), perceived general level of knowledge (n=6), and high-risk groups (n=5). The level of knowledge for the different variables was calculated using scores (high or low scores) or only reported in the percentage of participants. Several studies reported only an overall knowledge score or a proportion of respondents with a high level of knowledge (generally at least 70% correct answers), without reporting scores for each knowledge variable measured (e.g., symptoms, modes of transmission) (23, 38, 39).

##### General adult populations

Adult participants had an overall good knowledge of COVID-19. In most studies, the overall knowledge rates and the proportions of knowledgeable respondents were relatively high (from 72% to 98% of respondents) (22, 26, 29, 37, 38). In two studies, however, the survey results showed that respondents were not very knowledgeable: a mean score of 8.56/13 among Bangladeshi respondents (39) and 41% of the respondents having poor knowledge among the adults in the United States (23). In addition, three studies in Hong Kong (24), India (27), and Sudan (37) found that a significant proportion of individuals (ranging from 24% to 38%) were not aware that asymptomatic persons can infect others/or know the period of asymptomatic incubation. Similarly, a very high proportion of respondents (>90%) identified that the disease could be transmitted through droplets, and direct or indirect contact (between 99% and 83%) (24, 26, 28), except in one study where only 29% of the Indian respondents knew that COVID-19 spreads through multiple modes like touching, kissing, and sneezing (25). Adult respondents were also very knowledgeable about the common symptoms of COVID-19. In several studies, a minimum of 80% of the respondents (up to 98%) knew all of the common symptoms of COVID-19 (26, 28, 30, 37), except in one study conducted among Indian adults where only 18% of the respondents considered fever to be a symptom of the disease (25). Finally, adult participants were very knowledgeable about preventive practices to avoid COVID-19 transmission, with more than 90% of the respondents acknowledging the preventive behaviors like social distancing, hand washing/sanitizing, wearing a mask, and avoiding public gatherings (25-27).

##### High-risk adults

Studies focusing on high-risk adults also found that most respondents were very knowledgeable about COVID-19, including common symptoms, routes of transmission, and behaviors to avoid an infection (proportions of respondents were between 77% and 94%) (32-34, 36, 41). As for adults in general populations, one study found that only 26% of liver recipients and candidates for transplants thought that COVID-19 can be spread by asymptomatic patients (41). The two studies conducted with poor households also showed a lack of knowledge about difficulty in breathing being a common symptom of COVID-19 (36) and some preventive behaviors such as social distancing, wearing a mask, and avoiding crowded places (32). Finally, the study conducted with pregnant women revealed an important lack of knowledge regarding the impact of COVID-19 on preterm births (35).

#### Preventive behaviors towards COVID-19

Several preventive behaviors were assessed in the 20 included studies, including hand-washing (n=15), wearing a mask (n=15), staying at home/reducing social contacts (n=13), avoidance behaviors (e.g., avoiding crowded places, social gatherings or public transports, cancelling travel) (n=11), and practicing social distancing (n=8).

##### General adult populations

In studies on general populations, many authors reported appropriate behaviors for preventing COVID-19. The most observed preventive behavior was washing hands frequently, with reported rates from 68% to 99% among respondents (24-30, 37, 38). According to several studies, avoiding crowded places or social gatherings were also practices generally adopted by most respondents (from 65% to 99%) (25, 27, 37-39), except in one study where a minority of South Korean participants (41%) reported avoiding crowded places (29). In any case, adults were reportedly more or less compliant when it comes to staying at home, reducing social contacts, or avoiding public transport. In three studies, more than 80% of respondents followed these practices (22, 25, 28, 38) whereas studies conducted among Hong Kong and South Korean adults showed lower rates in adopting preventive behaviors; 53% and 39% of the respondents, respectively, reported avoiding public transport (24, 29). Mask wearing was also variably followed in the studies. While five studies reported high proportions of respondents (63% to 97%) wearing masks (24, 27, 29, 38, 39), four other studies conducted among Egyptian, Serbian, Sudanese, and

Indian adults showed much lower compliance (around 35%) (25, 26, 30, 37). Finally, practicing social distancing was also variably followed. For example, studies conducted among Ugandans and Serbians showed more compliance among the participants with this practice (30, 38) compared to the respondents in South Korea (29).

##### High-risk adults

Overall, studies on the high-risk adults have reported appropriate preventive behaviors during the early period of the pandemic. Two studies reported that a large majority of respondents (>90%) among poor households, liver recipients, and candidates for transplants washed their hands more frequently (34, 36). Nevertheless, only 40% of the adults with Parkinson’s disease reported washing their hands more frequently (40). Staying at home or leaving home less frequently were also reported by a majority of respondents (60-79%) from poor households (36), liver transplant recipients, candidates for transplants (34, 41), and people with Parkinson’s disease (40). Most of the respondents (63%-94%) from poor households (32, 36) and sexual minorities (33) avoided crowded places or stopped attending social gatherings. In three studies involving liver transplantation recipients and candidates for transplants in India (41), poor households (32) and adults with Parkinson’s disease (40) few participants reported wearing a mask (6%-40%). Nevertheless, in Germany, 78% of the liver transplantation recipients and candidates reported wearing a mask when leaving home (34). Finally, the two studies conducted among poor households in Kenya (36) and the Philippines (32) reported a high proportion of respondents (81% and 66%, respectively) keeping distance from other people to avoid getting infected from COVID-19.

#### Factors associated with RPKB towards COVID-19

The most studied factors that were significantly associated with RPKB towards COVID-19 were socio-demographic factors such as age, gender, education, and ethnicity.

##### Age

Studies in Hong Kong (24) and South Korea (29) found that older adults were significantly less worried about getting infected compared to the young adults. We found one exception to this in a study among poor households in Kenya where the perception of risk increased by age group (36). Age was positively associated with perceived severity in two studies: one in Serbia among the public (30) and the other in the United States among persons with chronic conditions (31). In four studies, older adults were also found to be more knowledgeable about COVID-19 compared to younger adults (28, 30, 34, 39). In addition, three studies found that age was positively associated with hand-washing (28, 29, 37).

##### Gender

In two studies, women were significantly more worried about being infected with SARS-CoV-2 or to consider COVID-19 as a threat to health (31, 34, 40). Men were found to be less knowledgeable than women about COVID-19 in three studies (28, 30, 37). Finally, a high number of studies reported that the adoption of preventive behaviors such as hand-washing, wearing a mask, reducing contacts, avoiding social gatherings, or practicing social distancing was positively associated with the female gender (22-24, 28, 29, 31, 37,39).

##### Level of education

A higher level of education was positively associated with knowledge of COVID-19 in five studies, especially with regards to modes of transmission and common symptoms (24, 30, 32, 36, 37). Adults who were educated were also more likely to adopt preventive behaviors like wearing a mask or maintaining social distance (29, 30, 32).

##### Ethnicity

Significant differences were found in RPs between ethnic groups in two studies that reported that Black respondents were less likely to be worried about getting COVID-19 (23, 31). In three studies, the Black respondents were also less likely than the White respondents to have a high knowledge of COVID-19 (23, 28, 31). Finally, some studies showed mixed evidence regarding ethnic disparities in adopting preventive behaviors. One study with a low-quality rating was conducted in the general population. In this study, Black people were reported to be more likely to have good practices towards the transmission of COVID-19 (23). Another study, with a medium-quality rating and involving patients with chronic conditions, showed the opposite results (31).

## Discussion

To the best of our knowledge, this is the first scoping review that offers a mapping of studies conducted among general and high-risk adult populations on RPKB towards COVID-19 and factors associated with RPKB.

Our scoping review has several limitations. We decided to cover a short period (January-June 2020). In the context of the pandemic, a large number of studies were conducted and published very early. Consequently, a first step in mapping the emerging evidence has been to gain an understanding of RPKB towards COVID-19 in the early stage of the pandemic. We did not provide a cross-comparison between countries, mainly because of the significant heterogeneity of studies regarding survey and sampling methods and the absence of cross-country studies in the scoping review. Finally, because the overall quality of the included articles was quite low, the research findings presented here should be interpreted carefully. Most studies used non-probability sampling and online surveys that raises doubts about the capacity for authors to generalize the research findings. While online surveys allow a rapid and user-friendly data collection from large samples of the population, they can also increase the likelihood of sampling and non-response bias (42).

Overall, in the early months of the pandemic, the levels of RPKB towards COVID-19 were moderate to high in both general and high-risk populations. We did not notice significant differences in RPs between the general and the high-risk adult populations. Nevertheless, two studies, one with pregnant women in Turkey (35) and the other with poor households in Kenya (36) reported low-risk perception levels, in contrast to six other studies conducted among high-risk adults (31-34, 40, 41). Interestingly, overall, the perceived severity of the disease was slightly higher than the perceived susceptibility of getting COVID-19 during the first months of the pandemic. Similar findings were reported in an international study by Zwart et al. (43) on RPs related to SARS-CoV, which revealed an intermediate level of SARS vulnerability and a high perceived severity, in comparison to other diseases (43).

This finding might seem counterintuitive since we now know that the case-fatality rates for COVID-19 are relatively low while the transmissibility rates are high, in comparison to other coronavirus disease outbreaks (44). Nevertheless, several explanations could be given for this finding. During the first wave of the COVID-19 pandemic, a delay occurred before a scientific consensus on mortality rates and asymptomatic transmission emerged and before the public was informed. Another explanation could be that during the first months of the pandemic, cases of COVID-19 were highly concentrated in certain regions (e.g., Hubei in China, Lombardy in Italy) or cities (e.g., Wuhan, Milan, New York City) (45, 46), which providing a false sense of security towards COVID-19 transmission for people living outside the COVID-19 hotspots. This difference in risk perception between hotspots and safe-zones agrees with a study conducted in China during the early stage of COVID-19. Shanghai respondents were reported to have significantly lower perceived susceptibility and higher severity, compared to their counterparts in Wuhan (47). Finally, the anxiety caused by the media and the memory of past fatal outbreaks, such as those caused by MERS and Ebola could explain that people were more worried about dying from the disease than being infected at the very beginning of the pandemic. This explanation is in line with Zwart et al.’s findings on RPs during the SARS outbreak that indicated that more unfamiliar diseases can be perceived as being more severe (43).

The scoping review showed that general and high-risk adults were knowledgeable about COVID-19. Our findings are consistent with those of Majid et al.’s recent scoping review on knowledge, RPs, and behavior change during pandemics, where the authors stated that knowledge generally spreads rapidly during pandemics in most regions (48). Nevertheless, we found exceptions in several studies that reported a relatively low level of overall knowledge among the general public in Bangladesh (39), and the United States (23). Overall, the participants were very knowledgeable about preventive behaviors, including hand-washing, mask-wearing, social distancing, and avoidance behaviors. Nevertheless, an important knowledge gap on the asymptomatic transmission of COVID-19 was reported in many studies (24, 27, 37) as the asymptomatic nature of the virus transmission had not been clearly scientifically identified or shared with the public during the first wave of the pandemic. A high proportion of the respondents from poor households in Kenya did not identify difficulty in breathing as one of the main symptoms of COVID-19 (36). Similarly, the pregnant women participants in Turkey were unaware that COVID-19 can cause preterm births (35), though this study was evaluated to be of low quality. Conversely, Maharlouei et al. (49) found that a greater proportion of pregnant women were aware of the risk of severe complications during birth.

Our review identified hand-washing and avoiding crowded places as dominant preventive behaviors among both general and high-risk adults at the early stage of the pandemic. Nevertheless, staying at home, reducing social contacts, and avoiding public transport were less widespread in general populations. Alternatively, the high-risk adults reported being much more compliant with staying at home or leaving home less frequently (34, 36, 41). Wearing a mask was the least respected practice in the early stage of the pandemic for both general and high-risk adults, except in one study conducted in Hong Kong where 97% of the participants reported wearing a mask when leaving home (24). In Majid et al.’s scoping review, the authors reported varying degrees of adopting mask wearing, ranging from 4% in the United States to 96% in China (48). In East Asia, mask wearing is socially embedded as a general preventive practice (50). Surprisingly, a large majority of the participants from poor households (32, 36), including those living in slums in Kenya (36) reported using social distancing to avoid getting infected. This finding is in contradiction with a recent observational study conducted in an urban slum in India. In any case, the authors concluded that social distancing measures were more of an aspiration than reality (51).

Our review highlighted the existence of significant sociodemographic differences in RPKB towards COVID-19. Being a female, older, and more educated was associated with better knowledge about COVID-19 and appropriate preventive behaviors. This finding is consistent with Bish and Michie’s review on the determinants of protective behaviors during a pandemic (8). We also found that age was negatively associated with perceived susceptibility to and positively associated with perceived severity of the disease. In a recent study on the age effect on preventive behaviors during the COVID-19 pandemic, the authors concluded that the oldest adults underestimated the probability of getting infected but at the same time they were aware of the COVID-19 threat (52). We also found several studies that showed Black respondents being less worried about getting infected and less knowledgeable about COVID-19, compared to White respondents (23, 28, 31). At first glance, we might hypothesize that lower RPs and knowledge about COVID-19 among Black individuals might lead them to be less compliant with preventive behaviors. Nevertheless, mixed evidence has been given for the link between ethnicity and adoption of preventive behaviors, a finding that is consistent with the review of Bish and Michie (8).

## Conclusion and future research

Our findings have several implications for public health authorities aiming to be responsive in adopting appropriate and effective risk communication strategies at the very early stages of a pandemic. Our review showed that during the first wave of COVID-19, the perceived severity of COVID-19 was higher than the perceived susceptibility among general and high-risk adult populations. This finding suggests that people, especially those in less-affected areas, might have underestimated the infectivity of the virus. Perceived susceptibility combined with perceived severity plays a vital role in motivating health protection behaviors (53) and may facilitate or reduce the transmission of a virus during pandemics (11, 14). While countries are facing a second or third wave of COVID-19, RPs should be continuously monitored to adjust the risk communication strategies over time. Effective risk communication relies on generating a sense of worry among the public while avoiding the fear that could lead to denial and inappropriate behaviors (54). Communication strategies must target certain groups of the population, including men, young and less educated adults who are less likely to perceive the risks associated with COVID-19, and those who are less knowledgeable and less likely to adopt preventive behaviors. Addressing literacy and numeracy issues in less-educated people can be achieved by delivering simple communications using videos about the preventive behaviors (55). Young adults can be targeted by using accessible, credible, and reliable social media channels for providing information about COVID-19 (56).

While this review offers an initial understanding of RPKB of adult populations during the early stage of the COVID-19 pandemic, further research is needed to assess the psychological and behavioral responses over time. Whether people from ethnic minorities are less or more likely to have high levels of preventive behaviors towards COVID-19 is unknown. In any case, the disproportionate number of COVID-19 fatalities within the Black population (4, 6) should alert us to possible gaps in RPKB towards COVID-19 in these communities. Additional studies on ethnic health disparities may help public health authorities to introduce targeted actions towards these communities during the COVID-19 pandemic. While population-based surveys allow for a rapid assessment of psychological and behavioral responses of populations, in-depth qualitative studies are necessary to acquire a deeper understanding of RPKB towards COVID-19 among high-risk groups in the population.

## Supporting information

Supplementary file 1 - Medline-Ovid search strategy

Supplementary file 2- Quality appraisal

## Data Availability

All data referred to in the manuscript are available through the link below.

https://www.researchgate.net/publication/348663541_Risk_Perceptions_Knowledge_and_Behaviors_of_General_and_High-Risk_Adult_Populations_towards_COVID-19_A_Systematic_Scoping_Review

