## Supplementary file 1 - Medline-Ovid search strategy for "Risk Perceptions, Knowledge and Behaviors of General and High-Risk Adult Populations towards COVID-19: A Systematic Scoping Review"

### Appendix 1: Search strategy in Ovid-MEDLINE

1. exp coronavirus infections/
2. (betacoronavirus* or coronavirus* or corona virus*). mp
3. 1 or 2
4. exp china/
5. (china or chinese or hubei or wuhan).af.
6. 4 or 5
7. 3 and 6
8. (betacoronavirus* or coronavirus* or corona virus*).ti,kf.
9. Severe acute respiratory syndrome coronavirus 2 or "SARS CoV-2" or "SARSCoV 2" or SARSCoV2 or cov2 or "sars 2" or COVID or "coronavirus 2" or covid19 or ncov or ((novel or new) adj3 coronavirus) or ncp). mp.
10. 8 or 9
11. Exp pneumonia/
12. Pneumonia.mp.
13. 11 or 12
14. Wuhan.af.
15. 13 or 14
16. 7 or 10 or 15
17. (percept* or perceiv* or representat* or aware* or conscious*).kf,tw.
18. (knowledge or understand* or comprehens*). kf.tw.
19. (behavior* or behaviour* or action* or attitude* or practic*).kf,tw.
20. (population* or person* or people or invidual* or adult* or citizen* or resident* or public or communit* or group*). kf,tw.
21. 17 or 18 or 19
22. 16 and 20 and 21
23. Limit to 22 (english language and humans and yr="2020-current")

The 23 step MEDLINE strategy yielded 2439 references.
