## Supplementary file 2- Quality appraisal for "Risk Perceptions, Knowledge and Behaviors of General and High-Risk Adult Populations towards COVID-19: A Systematic Scoping Review"

### Appendix 2: Quality appraisal of the included studies

| **Quality assessment criteria (AXIS tool)** | 1. Clear study objectives | 2. study design appropriate | 3. Sample size justified | 4. target population clearly defined | 5. sample represents the target population | 6. selection process appropriate (sample) | 7. Measures undertaken to adresss non-response rate | 8. Outcome variables appropriate | 9.Measurements validated/piloted or published | 10. Statistical significance clearly presented | 11. Methods sufficiently described | 12. Basic data adequately described | 13. Response rate does not raise concerns about non-response bias | 14. Non-responders described | 15. Results internally consistent | 16. results from all analyses described | 17. Discussions and conclusions justified by results | 18. Limitations discussed | 19. Absence of conflict of interests | 20. Ethical approval and participant consent | Total score (/20) | **Total score (%)** |
|---|---|---|---|---|---|---|---|---|---|---|---|---|---|---|---|---|---|---|---|---|---|---|
| Abdelhafiz AS, Mohammed Z, et al | yes | yes | yes | yes | no | not reported | not reported | yes | yes | yes | no | no | no | no | yes | yes | yes | no | yes | yes | 12/20 | **60** |
| Alobuia WM, Dalva-Baird NP, et al | yes | yes | no | yes | not reported | not reported | not reported | not reported | no | yes | no | yes | no | no | no | yes | yes | yes | not reported | not reported | 8/20 | **40** |
| Alsan M, Stantcheva S, et al | yes | yes | no | yes | yes | yes | no | yes | not reported | yes | yes | yes | no | no | yes | yes | yes | yes | yes | yes | 15/20 | **75** |
| Austrian K, Pinchoff J et al | yes | yes | yes | yes | no | no | not reported | yes | yes | yes | no | no | yes | no | yes | no | yes | yes | not reported | not reported | 11/20 | **55** |
| Bostan S, Erdem R, et al | yes | yes | no | yes | no | not reported | not reported | yes | yes | yes | no | no | no | no | no | yes | no | no | not reported | not reported | 7/20 | **35** |
| Chan EYY, Huang Z, et al | yes | yes | yes | yes | yes | yes | no | yes | yes | yes | yes | yes | no | no | yes | yes | yes | yes | yes | yes | 17/20 | **85** |
| Cvetković VM, Nikolić N, et al | yes | yes | no | yes | no | not reported | not reported | yes | yes | yes | yes | no | no | no | no | yes | yes | yes | yes | yes | 12/20 | **60** |
| Hakeem AR, Padmanaban H, et al | yes | yes | yes | yes | yes | yes | yes | yes | no | not relevant | yes | yes | yes | yes | yes | yes | yes | no | yes | no | 17/19 | **89** |
| Hezima A, Aljafari A, et al | yes | yes | yes | no | no | no | not reported | yes | no | yes | no | yes | no | no | yes | no | yes | no | yes | not reported | 9/20 | **45** |
| Ko N-Y, Lu W-H, et al | yes | yes | no | yes | no | not reported | not reported | yes | yes | yes | yes | no | no | no | yes | yes | yes | yes | yes | yes | 13/20 | **65** |
| Lau LL, Hung N, et al. | yes | yes | yes | yes | no | no | not reported | yes | yes | yes | yes | yes | yes | no | yes | yes | yes | yes | yes | yes | 16/20 | **80** |
| Lee M, You M | yes | yes | no | yes | no | not reported | not reported | yes | yes | yes | yes | no | no | no | yes | yes | yes | yes | not reported | yes | 12/20 | **60** |
| Parikh PA, Shah BV, et al | yes | yes | no | yes | no | no | not reported | yes | no | not relevant | no | no | not reported | no | no | yes | yes | yes | yes | yes | 10/19 | **53** |
| Paul A, Sikdar D, et al | yes | yes | no | yes | no | no | not reported | yes | yes | yes | yes | no | not reported | no | yes | yes | yes | no | not reported | yes | 11/20 | **55** |
| Reuken PA, Rauchfuss F, et al | yes | yes | no | yes | not reported | not reported | not reported | yes | no | yes | yes | yes | not reported | no | yes | yes | yes | yes | yes | yes | 13/20 | **65** |
| Roy D, Tripathy S, et al | yes | yes | no | no | no | no | not reported | yes | no | not relevant | no | no | no | no | yes | yes | yes | yes | yes | yes | 10/19 | **53** |
| Ssebuufu R, Sikakulya FK et al | yes | yes | no | yes | no | no | no | yes | no | yes | yes | no | no | not reported | yes | yes | yes | yes | not reported | yes | 11/20 | **55** |
| Wolf MS, Serper M, et al | yes | yes | yes | yes | no | no | no | yes | no | yes | yes | yes | yes | no | yes | yes | yes | yes | yes | yes | 15/20 | **75** |
| Yassa M, Birol P, et al | yes | yes | no | yes | not reported | no | not reported | yes | no | not relevant | yes | no | not reported | no | no | yes | yes | no | yes | yes | 10/19 | **53** |
| Zipprich HM, Teschner U, et al | yes | no | no | yes | not reported | not reported | not reported | yes | no | yes | yes | yes | yes | no | no | yes | yes | yes | yes | yes | 12/20 | **60** |
| **Mean score (%)** | | | | | | | | | | | | | | | | | | | | | | **61** |
